# Over-dispersion in Malaria–Schistosoma Co-infection: Insights from a Meta-analytical Approach and Systematic Review

**DOI:** 10.1101/2025.10.31.25339164

**Authors:** Célia Koellsch–Amet, Jérôme Boissier, Ronaldo de Carvalho Augusto

## Abstract

Malaria and schistosomiasis are two major parasitic diseases that are co-endemic in many regions of sub-Saharan Africa. Despite their frequent overlap, the potential epidemiological interactions in cases of co-infection remain poorly understood. We conducted a systematic review and meta-analysis in accordance with the PRISMA-ScR guidelines. A total of 192 studies published between 1996 and 2023 were reviewed, of which 59 studies involving 73,383 individuals were included in the meta-analysis. Pooled analysis showed that *Plasmodium– Schistosoma* co-infection occurs more often than expected by chance (overall odds ratio [OR] = 1.20, 95% CI: 1.02–1.40), despite substantial heterogeneity (I^2^ = 89%). Co-infection prevalence ranged from 1.1% (Benin) to 36.6% (Mali), with school-age children and pregnant women disproportionately affected. Subgroup analyses revealed no consistent differences by sex or by *Schistosoma* or *Plasmodium* species. Observational and experimental evidence suggests that co-infection may exacerbate anemia and modulate host immunity, but mechanistic pathways remain poorly defined. The high co-endemicity of these parasites underscores the need for integrated surveillance and control programs and highlights the potential interactions between the two parasites. Understanding these mechanisms is essential for designing integrated control strategies and highlights the need to take polyparasitism into account in public health policies, particularly in endemic regions.

## Introduction

Malaria and schistosomiasis are among the most widespread and debiliting parasitic diseases affecting 85 and 78 countries, respectively^1,2^. On a global scale, it is estimated that 46.4 million DALYs (Disability-Adjusted Life Years) was lost due to malaria ^3^, compared with 1.6 million for schistosomiasis ^4^, making these two parasitic diseases the two most important in the world; More than a third of the world’s population is thought to be infected by helminths or *Plasmodium* ^5^ with Africa remaining the hardest-hit continent. Sub-Saharan Africa bears the greatest burden of both infections, where overlapping ecological and socio-economic conditions create extensive zones of co-endemicity^6–10^. Moreover, the epidemiology of co-infection is influenced by a variety of factors including population dynamics, behavioural, genetic, host physio-immunology, population size, and parasite dispersal patterns^6^. These include the high frequency of the two parasites in the same population, the similar geographical distribution of the vectors and the need for an aquatic environment to complete both parasite life cycles. Whether plasmodium or schistosome, the two parasites have complex life cycles, dependent on environmental conditions that allow other host species - mosquitoes for *Plasmodium*, freshwater snails for *Schistosoma* - to thrive. Furthermore, spatial statistical models highlight the geographical overlap and thus support the co-endemicity of malaria and schistosomiasis infections ^11^. Environmental sensitivity and underprivileged socio-economic conditions have repeatedly been reported to influence the survival of free-living invertebrate hosts and helminths, thereby governing the spatial distribution of diseases and favouring the geographical overlap of *Plasmodium* and *Schistosoma*^7,12,13^.

Since the last major pandemic of COVID-19 that recently affected the world, the question of an interaction between climate change and infectious diseases has become increasingly worrying ^14^. As the effects of these changes are still poorly understood, climate change as a whole is generating major uncertainties about the epidemiology of many diseases, and is widening the gap between current healthcare techniques and the evolution of the epidemiology of infectious diseases. To date, it is estimated that 58% of infectious diseases in humans have at some time been intensified by climatic events^15^. Climatic factors can also lead to the emergence of new pathogens and increase the risk of transmission of existing diseases such as malaria and other neglected tropical and subtropical diseases such as schistosomiasis ^16–18^. In Africa, malaria episodes are impacted by environmental variability and often occur after climatic anomalies such as periods of drought or extreme rainfall ^19,20^. However, falciparum malaria remains confined to tropical and subtropical regions in the various predictive models ^21^, while *Plasmodium vivax*, formerly the dominant species in Europe, could re-emerge ^22,23^. An outbreak of schistosomiasis has also recently been identified in southern Europe (Corsica, France), with infected patients originated from France, Germany and Italy ^24,25^, resulting in both of an increase in the human migration and of the rise in the temperature of fresh water ecosystems, which favours the establishment of the snail vector in southern Europe^26^. Identifying current hotspots of co-endemicity and understanding the ecological drivers of overlap will be critical for anticipating future shifts in disease distribution.Although the epidemiology and immunology of major parasitic diseases such as malaria and schistosomiasis are relatively well documented, the mechanisms and implications of concomitant infections remain largely unexplored. However, co-infection with *Plasmodium spp*. and *Schistosoma spp*. is common in many endemic regions and could be modulated by global warming, prompting growing interest in understanding the extent to which these parasites can interact in the same host ^18,27^. In particular, several studies suggest that *Schistosoma* infections may modulate susceptibility or immune response to *Plasmodium* infection, raising key questions about the potential interactions between these pathogens, both clinically and immunologically. Interactions between *Plasmodium* and helminths have already been demonstrated to influence the host immune response to malaria infection ^28,29^. In the specific case of schistosomiasis, several studies also suggest the existence of cross-effects with malaria, although the exact nature of these interactions - protective or deleterious - remains controversial ^30–39^. Some studies report that co-infection with schistosomes could worsen *Plasmodium* infection, by increasing the intensity, prevalence or clinical impact of *Plasmodium spp*. ^30,33,37,40,41^. Conversely, other studies indicate a mitigating or even protective effect of schistosomiasis on the severity of malaria ^31,36,39,41^. At the same time, some studies have found no significant association between these two infections ^42–45^. These controversies may stem from the fact that sampling protocols vary between studies. The strains of parasites studied may also differ from one study to another, as may the sampling region. The timing of infection could influence the outcome of co-infection, whether simultaneous or sequential and this still is an aspect neglected in epidemiological analyses.

Observational and experimental studies, in both animals and humans, have begun to reveal the mechanisms of these interactions, whether direct (through competition for resources) or indirect (via modulation of the immune response). Nevertheless, the complexity of interactions between *Plasmodium spp*. and *Schistosoma spp*. continues to be investigated, with the aim of better defining the synergistic or antagonistic effects of these co-infections in human populations^46–48^. To date, the appearance of simultaneous infections by these two parasites in humans seems to be at the origin of the emergence of several studies aimed at elucidating aspects related to the synergistic and antagonistic interactions of *Plasmodium spp*. and *Schistosoma spp*.^46–48^.

Despite the reported effects of malaria and schistosomiasis infections, the nature of the interactions between the two parasites remains unclear. It is possible that this is due to the general complexity of the interactions between parasites and of the pathways involved in these interactions during co-infection. Thus, a clear understanding of the epidemiology of malaria during co-infection with schistosomiasis is essential to inform decisions on appropriate control strategies against these two diseases. This systematic review and meta-analysis provide, a review of the epidemiological and experimental studies identified on *Schistosoma* and *Plasmodium* interactions and interprets the plausibility of these interactions. Consistent with the studies conducted, we hypothesize that *Plasmodium* and *Schistosoma* interact positively during co-infection and generate both an impact in their host.

## Method

### Protocol and registration

This systematic review and meta-analysis was conducted in accordance with the recommendations of the PRISMA-ScR 2020 guidelines (Preferred Reporting Items for Systematic Reviews and Meta-Analyses extension for Scoping Reviews), as detailed in appendix S1. The aim of the approach was to explore and synthesise the available data on interactions between Plasmodium *spp*. and *Schistosoma spp*. in contexts of human co-infection.

### Information sources and search strategy

A systematic search was conducted in two bibliographic databases: BibCNRS and PubMed, on 24 January 2025. The search strategy combined the terms “Plasmodium” OR “malaria” AND “Schistosoma” OR “schistosomiasis” AND “co-infection” OR “coinfection”. This search was completed by a citation search of articles included between 28 January and 6 March 2025.

Where article abstracts were not available, a selection was made from the tables of contents, followed by a full-text analysis. In addition, a manual grey literature search was conducted and managed using Microsoft Excel and Zotero software.

### Eligibility criteria

The studies included in this literature review met the following criteria: Epidemiological, experimental or functional studies reporting data on the prevalence or incidence of co-infection with *P. falciparum* or *P. vivax*, and *S. haematobium* or *S. mansoni;* Studies on general human populations, regardless of age or sex; Articles written in French or English and published in accessible scientific journals. Studies were excluded on the basis of the following criteria: Studies focusing solely on the treatment of infections; Unpublished studies, conference abstracts, protocols, non-accessible grey literature and articles in a language other than French or English; Studies mentioning co-infections with geohelminths without specific data on Schistosoma; Studies reporting no figures (prevalence, incidence, numbers, etc.) or not allowing a statistical effect to be calculated.

This analysis did not distinguish between asymptomatic and uncomplicated malaria, nor between the degrees of severity of the two parasitic diseases, in order to estimate the nature of the association between co-infection with Schistosoma and Plasmodium.

### Information and data extraction

The data were extracted into a dedicated Excel sheet, including the following information for each study: References (author, year); Period and location of study; Gender and age range of the population; Total number of participants and number of cases for each group (co-infected, mono-infected, uninfected); Prevalence rates for *Plasmodium spp*., *Schistosoma spp*. and co-infection; Additional information: anaemia, malnutrition, other associated pathologies.

These data were then imported into R via CSV files. The articles were classified by type: epidemiological surveys, functional studies and experimental studies.

Epidemiological studies are subjected to a meta-analysis, while other studies are subjected to a written synthesis. Using this spreadsheet, a preliminary summary of the results of the included studies was developed. Relationships within and between studies were also examined, as was the robustness of the synthesis for each study. The articles were grouped by type of study under three headings: epidemiological investigations, functional investigations and experimental studies. Each of these headings represents a paragraph of this review.

### Statistical analysis

Concerning epidemiological studies, in the Excel sheet of the included studies, the percentages of expected and observed co-infection were calculated (Appendix S2).

- Observed co-infection (actual co-infection measured in the study): This proportion is calculated from the number of people co-infected (infected with both Plasmodium and Schistosoma) in relation to the total sample size.

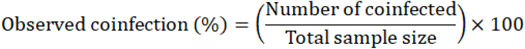
- Expected co-infection (if the infections were independent): This is a theoretical estimate of the co-infection that would occur if *Plasmodium* and *Schistosoma* infections were independently distributed in the population. It is calculated by multiplying the individual prevalence of each parasite:

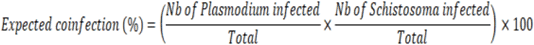

A Fisher’s test was performed to compare the expected and observed co-infection rates.The statistical analysis was then carried out in R using the metafor and meta packages. Meta-analyses were performed on the odds ratios (OR) using the Mantel-Haenszel method, with fixed-effects and random-effects models. Several sub-analyses were conducted: By type of co-infection (*S. mansoni* vs. *S. haematobium* / *P. falciparum* vs. *P*.*vivax*) ; By sex (men vs. women) ; By overall prevalence. An OR > 1 suggests that co-infection is more frequent than expected by chance and suggests a positive association and an OR < 1 suggests that co-infection is less frequent than expected, so a potential protective effect. Heterogeneity between studies was assessed using the I^2^ and Tau^2^ indicators. Forest plots were generated to visualise the results, and the presence of publication bias was analysed by funnel plot. This bias was also tested more formally using the Egger asymmetry test (metabias). A Fisher’s exact test was used to assess the association between certain categorical variables on a point-by-point basis.

### Co-occurrence map

To depict global patterns of malaria– schistosomiasis overlap, we compiled country-level data on malaria incidence and schistosomiasis occurrence from the World Health Organization Global Health Observatory and the Preventive Chemotherapy and Transmission Control (PCT) Databank (data accessed January 2025). Annual records from 2017–2023 were aggregated to classify each country as malaria only, schistosomiasis only, co-occurrence, or none. Maps were generated in R (v4.3) using the *ggplot2* and *choroplethr* packages.

## Results

### Search results

Database searches identified 2,064 articles (Figure 1). After removing duplicates, the titles and abstracts of 1,897 articles were reviewed, of which 105 articles were eligible for full review. A further 116 articles were identified by citation search, of which 112 were eligible. Of these 217 eligible articles, 25 were excluded for the following reasons (Figure 1): 18 did not evaluate the Schistosoma species that we were including in this review, 3 focused on drug treatments and 4 were neither in French nor in English. A total of 192 articles were included in the systematic review to construct the qualitative part of this review, of which 59 were used for the quantitative synthesis (meta-analysis) only. Interestingly, 34 articles were split between quantitative and qualitative analysis. As a result, the qualitative analysis (systematic review) alone included 167 articles. The characteristics of the articles included in the meta-analysis are summarised in Appendix S3.

**Figure 1.**
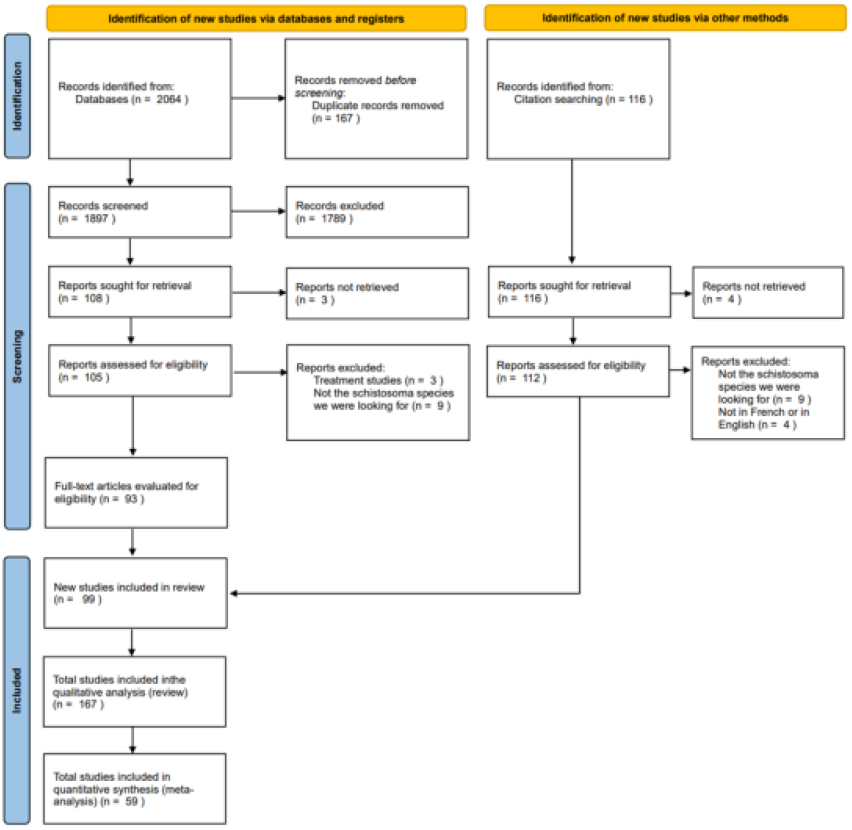
PRISMA 2020 flowchart of the process undertaken for the inclusion or exclusion of studies in the systematic review. PRISMA (Preferred Reporting Items for Systematic Reviews and Meta-Analyses) flowchart showing how 167 studies were obtained for the review. Of the 167 reports, 59 were included in the meta-analysis.

### Characteristics of the included studies

The characteristics of the 59 studies included in the meta-analysis are summarised in Appendix S3. All the studies were published between 1996 and 2023 and covered 14 countries - all on the African continent. Most studies were conducted in Nigeria (11/59, 18.64%). The selected studies included children and adults (both men and women). Co-infection between *S. haematobium* and *P. falciparum* were reported in 41 studies, 21 studies reported co-infection between *S. mansoni* and *P. falciparum*, 3 studies reported co-infection between *S. haematobium, S. mansoni* and *P. falciparum*, 1 study reported co-infection between *S. mansoni* and *P. vivax* and no study reported co-infection between *S. haematobium* and *P. vivax*.

#### 1. Epidemiology of co-infection

##### Spatial distribution of Plasmodium and Schistosoma

The epidemiological data (59 articles in total) covered the prevalence of co-infection and the morbidity associated with dual infection. Helminthic and malarial infections spread when climatic and environmental conditions are favourable to their development, particularly in low-income communities that require the installation of essential elements such as drinking water, water sanitation or improved hygiene of premises ^49,50^. In Tanzania, studies suggests linking the control of malaria-schistosomiasis co-infection to types of agro-ecosystem after demonstrating that children in rice-irrigated ecosystems were more likely to be co-infected than in other measured ecosystems ^9^. In countries such as Nigeria, Uganda, Ivory Coast and Ethiopia, helminth and malaria infections are highly prevalent and represent a persistent public health problem ^51–54^. More than a third of the world’s population, particularly those living in tropical and subtropical regions, are thought to have problems with concomitant infection by various species of Plasmodium and soil helminths ^6^.

Both malaria and schistosomiasis are widespread in similar tropical and subtropical areas, particularly in sub-Saharan Africa (Figure 2). Approximately 70% of the global burden of malaria is concentrated in sub-Saharan Africa ^55^ and 90% for schistosomiasis ^56^. These co-infections are particularly well studied in Nigeria and Uganda, where 11 and 9 studies respectively have investigated the prevalence of Plasmodium and Schistosoma co-infection (Supp.Fig.1). Similarly, the prevalence of co-infection was recorded for Ethiopia, Ghana and Ivory Coast in 6 studies each, for Mali in 5 studies, for Tanzania, Senegal, Kenya and Gabon in 4 papers, for Cameroon in 3 papers, for Zimbabwe in 2 papers and for the Democratic Republic of Congo and Benin in 1 paper each (Supp.Fig.1). Of the 59 articles considered for the epidemiological data, some analysed several countries. All the geographical areas listed in the studies analysed for this review are malaria- and schistosomiasis-endemic areas in sub-Saharan Africa.

**Figure 2.**
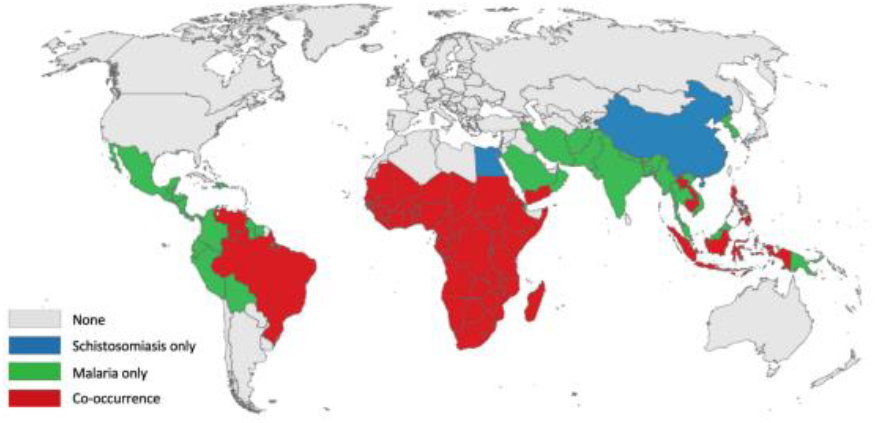
Global distribution of malaria, schistosomiasis, and their co-occurrence (2017–2022). Country shading indicates reported occurrence based on WHO Global Health Observatory data assessed in 2025: green – malaria only, blue – schistosomiasis only, red – co-occurrence of both infections, and grey – no reported occurrence.

##### Descriptive prevalence of malaria and schistosomiasis single and co-infections

Malaria is currently naturally caused by 9 species of Plasmodium: *P. falciparum, P. vivax, P. malariae, P. ovale curtisi, P. ovale wallikeri* found only in humans, as well as *P. knowlesi, P. simium, P. brasilianum* and *P. cynomolgi* which, although mainly found in monkeys, is also a pathogenic agent in humans ^57–59^. As far as schistosomiasis is concerned, there are currently more than 21 known species of Schistosoma, with 6 main species infecting Human (*S. haematobium, S. mansoni, S. intercalatum, S. japonicum, S. mekongi* and *S. malayensis*) ^60,61^.

For this meta-analysis, we have chosen to focus exclusively on infections with *P. falciparum* and *P. vivax as well as S. mansoni* and *S. haematobium* - the most widespread representative species from both parasites in Africa ^57,61^. The prevalence of *P. vivax* infection remains poorly studied in Africa despite consistent evidence of the emergence of this species on the continent (Supp.Fig.2A)^62,63^. The results of the studies clearly show the research interest in the infection prevalence of the other three species, with a maximum of 106 analyses carried out on the infection prevalence for *P. falciparum* in the 59 studies, compared with 86, 41 and 6 prevalence recorded for *S. haematobium, S. mansoni* and *P. vivax* respectively. The prevalence rates assessed in the various studies included in the meta-analysis were also higher for *S. mansoni* and *P. falciparum* species, with peaks of 92% and 100% respectively. The maximum prevalence analysed for *S. haematobium* and *P. vivax* were 78% and 22% respectively. Furthermore, unlike the other three species, the lowest prevalence of *P. vivax* was 13% (0% was observed for other parasites). The average prevalence of *S. mansoni, S. haematobium, P. falciparum* and *P. vivax* in these studies was 21%, 30%, 40% and 17% respectively on the African continent (Supp.Fig.2A).

In the present study, several types of co-infection were analysed, ranging from an association between two parasites to three. The most common and most studied co-infection was that between *P. falciparum* and *S. haematobium*, with 76 prevalence recorded and an average prevalence of 17.3%. This was followed by co-infections between *P. falciparum* and *S. mansoni* with 30 recorded prevalence and an average of 9.1%, between *P. falciparum, S. haematobium* and *S. mansoni* with 6 recorded prevalence and an average of 4.6%, between *P. vivax* and *S. mansoni* with 1 census and 0.9% prevalence and between *P. vivax, P. falciparum* and *S. mansoni* with 1 census and 11.2% prevalence (Supp.Fig.2B).

Differences in the prevalence of co-infections were found between the different African countries sampled in the meta-analysis studies. At the top of the list were Mali, with a prevalence of all co-infections combined of 36.6%, Nigeria with 25.8% and Gabon with 23.6%. Conversely, Benin, Ghana and Ethiopia had co-infections rates of less than 5% (Supp.Fig.3). However, there has only been one study of the prevalence of co-infection in Benin.

##### Meta-analysis of *Plasmodium spp*. and *Schistosoma spp*. Co-infection over-dispersion

Meta-analysis of the prevalence of co-infection in all studies combined, in cohorts composed of men, women and children, revealed statistical heterogeneity (I^2^: 89%; p-value < 0.0001) between studies (Figure 3). Overall, the prevalence of co-infections appeared to be over-dispersed, with a random effect not including the 1 (95% CI: 1.02; 1.40) (Figure 3). The most overdispersed estimated prevalences were found in a cohort from Senegal with an odds ratio of 39.86 (95% CI: 2.40; 661.19) and in second place in Kenya with an odds ratio of 15.40 (95% CI: 2.03; 117.04). The highest estimated prevalence of co-infection was 74%, found in a very small population of 54 individuals, while the lowest estimated prevalence was 0%, found in two studies of 404 and 6681 persons (Table S1). Two studies by Bassa et al. 2022 and Lyke et al. 2012 did not report the number of mono-infected patients who developed malaria (Table S1). However, one of the studies may be subject to sampling bias since the patients selected were all positive for malaria. (Table S1).

**Figure 3.**
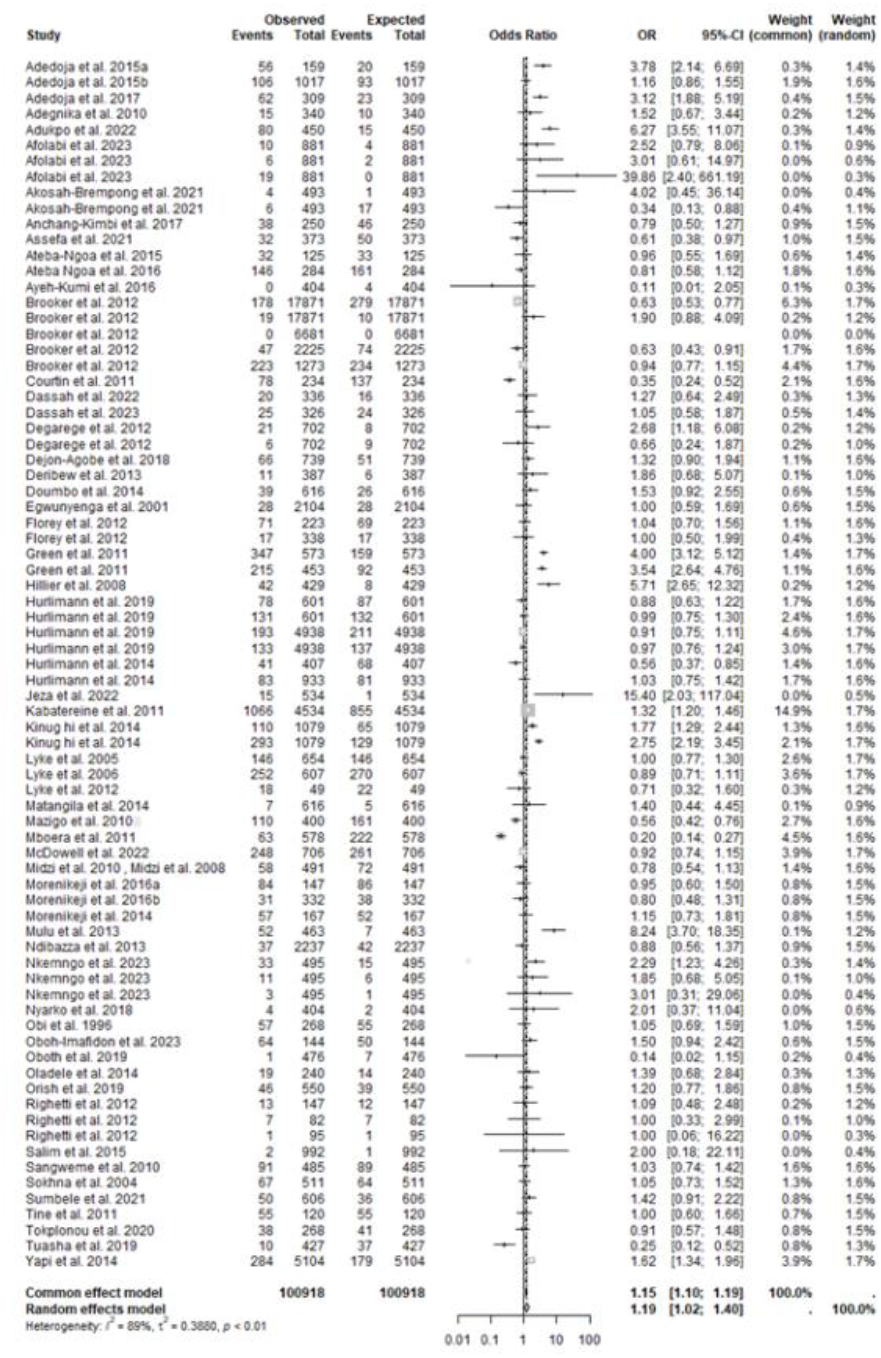
Forest diagram illustrating the results of the network meta-analysis for the overall prevalence of co-infections in studies that assessed the prevalence of malaria-schistosomiasis co-infections. Sensitive analysis comparing the difference between observed and expected co-infection. Blocks in the figure represent odds ratios, with error bars indicating 95% confidence intervals. Reports have been arranged alphabetically according to the first author. OR: odds ratio, CI: confidence interval, I^2^: heterogeneity coefficient, t^2^: inter-study variance.

##### Sex Stratification Reveals Consistent Over-Dispersion

A total of 73,383 men and women were sampled for the prevalence of co-infection across the 59 studies listed. Some studies distinguished between men and women in their sampling cohorts, while others did not. A sub-group analysis was therefore carried out, with women and men on one side (Figure 4). This highlighted the fact that, although a distinction is made between the two sexes in certain studies, there is no significant difference in the dispersion of the prevalence of co-infections between men and women (x^2^1 = 0.15; p-value = 0.70). There was statistical heterogeneity (I^2^: 48%; p-value = 0.0046), suggesting a moderate level of heterogeneity among the included studies for women, and statistical heterogeneity (I^2^: 27%; p-value = 0. 1430), suggesting a low level of heterogeneity among the included studies for men, although the Egger’s regression tests did not reach statistical significance (for women: Egger’s test = 1.83, p-value = 0.0805; for men: Egger’s test = 1.21, p-value = 0.2435). Overall, the meta-analysis by sex showed over-dispersion of co-infection in both 9726 women and 4593 men (Figure 4) in 22 and 15 studies respectively. This corroborates the results of the overall meta-analysis (Figure 3).

**Figure 4.**
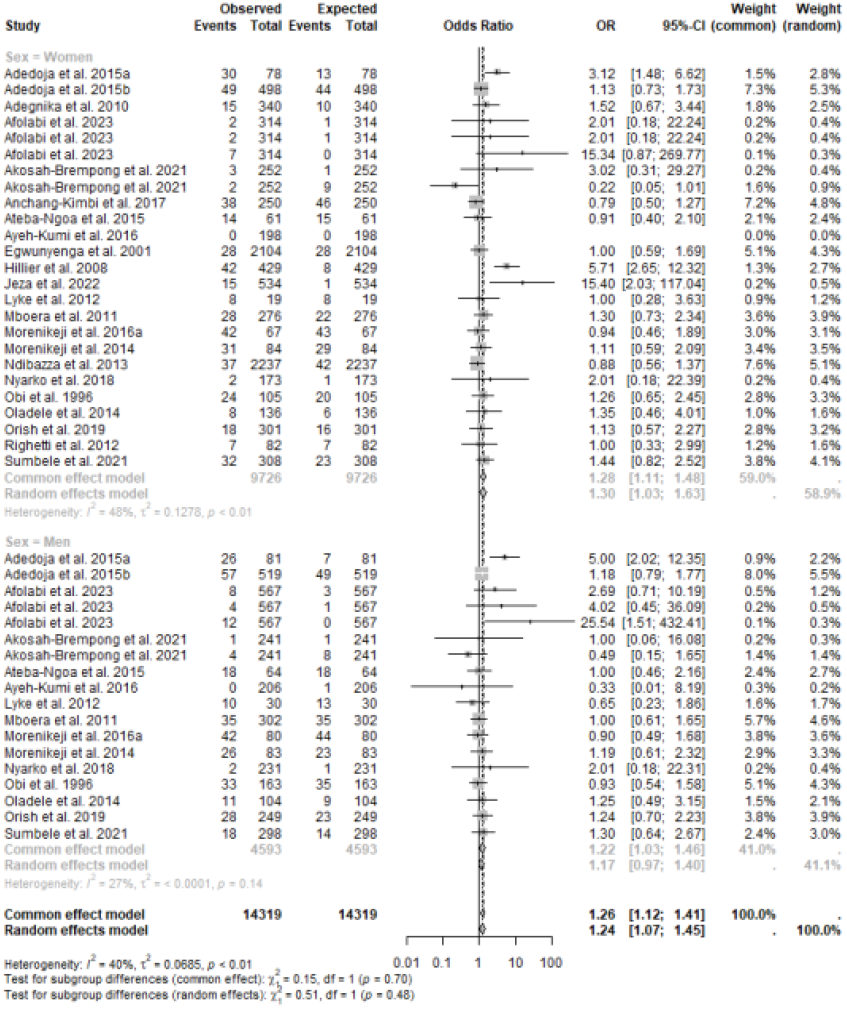
Forest diagram illustrating the results of the network meta-analysis for the overall prevalence of co-infections in women and men in studies that assessed the prevalence of malaria-schistosomiasis co-infections. Sensitive analysis comparing the difference between observed and expected co-infection in women and men. Blocks in the figure represent odds ratios, with error bars indicating 95% confidence intervals. Reports have been arranged alphabetically according to the first author. OR: odds ratio, CI: confidence interval, I^2^: heterogeneity coefficient, t^2^: inter-study variance.

We may envisage that the distinction between men and women, in the cohorts, was decided on the basis of socio-economic status and behaviour, which differ between communities but also between the two sexes. Indeed, demography and climatic factors, social and behavioural factors are associated with the risk of infection by the two parasites ^51^. Men are thought to be more exposed to the risk of schistosomiasis infection through long-term activities linked to water sources such as fishing or rice-growing ^64,65^ or simply bathing or washing in natural water sources ^66^. Moreover, in certain regions, for religious and socio-cultural reasons, women are sometimes forbidden to take part in activities such as swimming and fishing, which makes them less vulnerable to infection ^67,68^. Conversely, with regard to malaria infections, women would be more likely to be exposed to infections than men due to their prolonged exposure to mosquitoes during the most dangerous hours associated with outdoor domestic chores ^69^. However, this remains controversial, with several studies pointing to a greater risk of malaria infection in men^70,71^.

Co-infection rates of 46.8% were found among pregnant women in Cameroon, which can be explained mainly by the lack of water sources other than natural springs^72^. In contrast, co-infection prevalences of 23.8% and 20.8% were reported among pregnant women in Nigeria ^73,74^. This difference is mainly explained by the fact that pregnant women in some communities are denied access to natural water sources ^73^.

##### General clinical and immunological features of malaria and schistosomiasis co-infection

A major consequence of co-infection between malaria and schistosomiasis is anaemia ^29^. It is now known that these infections are both responsible for anaemia problems in humans, which could be exacerbated by the simultaneous presence of both parasites in the same host. For example, it has been shown that anaemia in Nigerien schoolchildren can be strongly influenced by a single infection with *Plasmodium spp*. or *S. haematobium*, but also by co-infection ^10^. Other studies have also found higher anaemia in co-infected patients compared to Plasmodium infection ^33,40,54,72,75–78^ (Table S2). It has also been shown that anaemia is greater in cases of co-infection than in cases of Schistosoma infection ^79^. However, other studies have reported contrasting results, showing that co-infection results in lower anemia levels compared to mono-infection with Plasmodium or Schistosoma ^10,45,80–82^ (Table S2). These discrepancies are hypothesized to arise from factors such as the genetic background of the human hosts and/or parasites, which may influence parasite-parasite and parasite-host interactions. In addition to the aforementioned data, co-infection with malaria and schistosomes is known to modify the immune profile of co-infected individuals, notably by altering the balance between the immune responses of TH1 response cells (helper T lymphocytes) and the TH2 response. Since Schistosoma induce a Th2 response in a chronic phase and a protozoan induces a Th1 response, co-infection of the two severely alters the development of an immune response and also affects already established responses^83^. Co-infection could also lead to a reduction in the immunological control of Plasmodium. Studies in humans and mice have shown that specific humoral and cell-mediated immune responses are essential for resistance to infection by both parasites^83^. Furthermore, murine malaria infection appears to significantly affect antibody levels in vivo and the cytokine response in vitro to *S. mansoni* antigen ^83^. Interestingly, *P. yoelii* infections reduced granuloma formation in the lungs of mice injected with *S. mansoni* eggs, indicating that malaria infection may influence granuloma formation in vivo ^84^.

Moreover, increasing evidence highlight the pathology seen in schistosomiasis and other parasitic diseases arises not only directly from parasite products but also from normal components of the immune response ^85^, in particular polymorphisms in cytokines such as IFN-γ, TNF-α, IL-4, IL-10, IL-13 and STAT-6 (Table S3). Some polymorphisms have been associated with susceptibility or resistance to Schistosoma infection ^86^. While other cytokine polymorphisms may be affiliated with the activity, amount and timing of cytokine production, influencing malaria susceptibility and severity ^87^. Given that *Plasmodium spp*. and *Schistosoma spp*. infections often coincide geographically in the same regions, it is crucial to know whether schistosomiasis infections modulate immune responses against the malaria parasite and affect its evolution.

#### 2. Coinfection of of Species Plasmodium and Schistosoma in experimental models

Several cases of co-infection with Plasmodium and Schistosoma in the murine model have already been tried, and numerous combinations have been tested: *S. mansoni* and *P. berghei* ^84,88–91^, *S. mansoni* and *P. yoelii* ^88,92–94^, *S. mansoni* and *P. chabaudi* ^83,88,95^ and *S. japonicum* and *P. berghei* ^96,97^. In addition, several of these studies have shown an effect of Schistosoma infection on the development of malaria, mainly reflected by an increase in parasitaemia ^83,93^ as well as an increase in the duration of malaria infection, resulting in the death of the mice. Conversely, the study by Waknine-Grinberg et al (2010) demonstrated that co-infection with *S. mansoni* and *P. berghei* in mice (not inbred) led to a change in Plasmodium infection and, in particular, increased survival in infected models. A change in cytokine expression due to the presence of Schistosoma was associated with a reduction in cerebral malaria.

However, co-infection with Schistosoma and Plasmodium is controversial in terms of its effects on either parasite. Indeed, some studies report a negative interaction between Plasmodium and Schistosoma ^39,47,96–98^ in which simultaneous infection with schistosomiasis and malaria resulted in lower densities of Plasmodium than in populations not infected with Schistosoma, suggesting that co-infection with schistosomiasis may have a protective effect against malaria ^99^. Although some studies demonstrate the possibility of a negative association resulting from coinfection, other studies have demonstrated an additive or synergistic effect of coinfection ^48,100^. In an earlier study in animal models, co-infection with schistosomiasis prolonged the time to low-density malaria parasitaemia and induced anaemia compared with the group infected with malaria alone ^101^. In many cases, however, it has been shown that previous infection with Schistosoma often has an effect on subsequent infection with a protozoan such as Plasmodium ^99^.

## Discussion

This systematic review with meta-analysis of 59 studies involving 73,383 individuals in 14 endemic African countries showed an overall mean prevalence of malaria-schistosomiasis co-infection of 13.41%, varying from 1.1% in Benin to an average of 36.6% in Mali. Helminth-malaria co-infection seems to be the subject of recent studies ^17,76,102,103^ and although polyparasitism is widespread in tropical and subtropical regions, its impact on public health has not been sufficiently studied to highlight the mechanisms involved. Our analysis, combining a co-endemicity map and the meta-analysis, suggests that co-infection between Plasmodium and Schistosoma is widespread in sub-Saharan Africa.

The overdispersion observed in *Plasmodium–Schistosoma* co-infections is probably explained by a complex set of biological, ecological and/or behavioural mechanisms. From an immunological and biological perspective, several hypotheses can be put forward. Co-infection may result from direct facilitation: a previous infection may alter the host’s immune response and facilitate the establishment of a second parasite, for example through immunomodulation or inappropriate polarisation of the response ^6,29^. Similarly, the cumulative effect of physiological stress caused by a first infection can weaken the host (loss of resources, alteration of the immune barrier), increasing the likelihood that another parasite will establish itself ^104–106^. This weakening could explain why some individuals infected with one parasite are more susceptible to a second infection. From an ecological point of view, the overdispersion of co-infection is largely influenced by the co-endemicity of the two parasites. Traditionally, co-endemicity and the overlap of the ecological niches of the two parasites are invoked to explain co-infection ^7,11^. Although this argument is more often used to justify coexistence rather than overdispersion, it can nevertheless contribute to it: indeed, repeated co-exposure linked to the ecology of vectors (mosquitoes and molluscs sharing the same habitats) can increase the probability of encountering both parasites simultaneously and reinforce their association within a part of the population. Climatic factors, such as soil moisture, rainfall and periods of drought, directly influence the survival and development of larval stages, thus determining the seasonality of transmission^29^. In a world subject to climate change, which is expected to increase habitat instability, parasites may face shorter periods of activity, which may force them to overlap in time. Finally, from the perspective of host-related factors, human behaviour is certainly a major determinant. School-age children are a particularly vulnerable population because they are more active, spend more time outdoors and are more likely to frequent contaminated water sources, which increases their likelihood of being infected by multiple parasites ^11,29^. Daily habits (swimming, washing clothes, fishing, irrigated agriculture) and the type of habitat (proximity to mosquito breeding areas or mollusc habitats) contribute to increasing this exposure. Beyond these behaviours, there is also intrinsic heterogeneity in host susceptibility: some individuals, known as ‘super-recipients’, are more susceptible to various infectious agents, either due to genetic characteristics, physiological or immune differences ^103,107,108^, or under the influence of gene-parasite interactions and environmental factors ^109,110^. These hosts account for a disproportionate number of co-infections and contribute significantly to the observed patterns of overdispersion ^111^. Genetic or physiological differences between individuals may also explain particular susceptibility profiles. Some hosts have genotypes that make their immune systems less effective against several infectious agents, contributing to a concentration of co-infections in a small fraction of the population ^107,112,113^. Finally, co-adaptation mechanisms between parasites have been suggested, whereby niche sharing could reflect an evolutionary strategy that limits direct competition in the long term ^108,111^.

In short, the over-dispersion of *Plasmodium–Schistosoma* co-infection cannot be attributed to a single factor. It results from a complex interaction between biological mechanisms (immunomodulation, fragility, genetic susceptibility), ecological factors (niche overlap, climatic influence, vector ecology) and host-related factors (risky behaviours, super-receptors). Understanding these interactions is essential for better predicting the dynamics of co-infections and refining control strategies in co-endemic areas.

Is important to highlight among the individuals analysed that it was common detect pregnant women co-infected by Plasmodium and schistosomes. It has been repeatedly demonstrated that pregnant women and young children are more likely to suffer high morbidity and mortality when infected with either malaria or schistosomiasis parasites ^73,105,114^. The importance of including pregnant women and young children in sampling cohorts therefore becomes clear when we understand that they are more likely to suffer the consequences of infection. Malaria-schistosomiasis co-infections are all the more important because of the more severe clinical symptoms and pathology than in the case of a mono-infection ^48,100,105^ and the possibility of modulation of the immune response ^106,115^ by the presence or interaction of the two parasites.

School-age children are also widely considered in co-infection studies because of their immune fragility and higher infection intensities compared with older individuals, the possibility of primary contact with in-utero infections that have made them fragile, and high-risk social behaviours such as bathing or washing in natural water sources ^43,116^. Indeed, in addition to the risk posed by the water feature in the transmission model for the two infections, children may be exposed to different points of infection when they go there - such as open defecation areas ^117^. Co-infections are also particularly common among school-age children in Nigeria, Uganda and Ghana ^44,50,53,75,76,118,119^.

In order to study the pathological and immunological mechanisms underlying this co-infection, the experimental model offers a number of advantages, providing access to more appropriate designs for the interaction and an ability to take advantage of the immune system of a predefined host - as in this case the mouse model, which benefits from a very well-characterised immune system. Although a few studies have used the simian model to study Plasmodium and Schistosoma infection/coinfection ^98,120^, the current understanding of the immunological basis of many disease processes was initially elucidated using mouse models, with studies of experimental *S. mansoni* infections in mice having been published since 1915 ^121–123^. All these denials demonstrate the importance of the experimental model for understanding the many aspects of coinfection on the virulence, compatibility and fitness of the parasite on several scales.

Despite the limited data available, which varies from one population to another, there is still no systematic and comparable information on the impact of polyparasitism among different age groups and in different epidemiological and nutritional contexts. Furthermore, much of the morbidity associated with polyparasitism remains unclear and poorly understood, such as the non-health-related societal consequences of cognitive impairment caused by helminthic and malaria co-infections ^124^. Anaemia is also a burden associated with schistosomiasis and malaria, and various studies have shown that the prevalence of anaemia can be significantly increased in cases of co-infection ^54,76,78^.

Research carried out in various epidemiological contexts has shown that polyparasitism occurs at intervals that are variable and dissimilar to those expected in the hypotheses of independence ^125–127^. Our meta-analysis confirmed an over-dispersion in the prevalence of co-infection compared with the expected results at the global level of the studies and between the sexes in the human host. There is increasing evidence that schistosomal infections are capable of altering susceptibility to clinical malaria ^31,35^ and there are now studies focusing on the mechanisms of co-infection, although these remain rare ^47,81^. In animal models, it has been suggested that these co-infections have both synergistic and antagonistic effects ^39,83,90,93,95^. Several hypotheses have therefore emerged to explain the interactions between Schistosoma and Plasmodium. Most studies on the interactions between helminths and malaria seem to indicate that helminth infection has a negative effect on the acquisition of immunity against malaria ^80,101,128^. It has been proven on several occasions that helminthic infection modulates the immune system of their host with the aim of survival^106^. One hypothesis is that helminths promote the production of non-cytophilic antibodies via the cytokine milieu, resulting in individuals who are more susceptible to clinical malaria infections ^29^. The presence of regulatory T cells intensifies during helminth infection, which can induce non-specific suppression by creating an anti-inflammatory environment ^129^. This non-specific suppression could extend to the immune response of other pathogens such as Plasmodium ^129^. Added to this is exposure to various environmental factors which, according to Smolen et al. 2014, could have an effect on the immune response of individuals. Indeed, this study compared the immune responses of children encountered on four different continents and demonstrated significant heterogeneity in innate cytokine responses in the different geographical areas sampled^110^. The differences in innate immune responses assessed were attributed to variations in environmental exposures such as feeding patterns, past infections, vaccination status, mode of delivery, region of residence, resource availability etc ^130^.

It has also been shown that the genetics of individuals represent a factor that can lead not only to infectivity but also to the severity of the disease in patients exposed to the same rates of infection ^112^. On the one hand, it seems that children born to mothers co-infected with Schistosoma and Plasmodium have a higher risk of malaria parasitaemia ^131^. Some studies have also shown that a fetus that has been exposed to maternal infections can react with hyporeactivity of the T lymphocytes that subsequently induce a reduction in immunity to these two parasites ^106,132^. On the other hand, the study by Oboh-Imafidon et al. 2023 revealed that people carrying a heterozygous (CT) or mutant (TT) CD14 (Cluster of Differentiation 14) genetic variant were up to 58% more likely to be co-infected than to be infected by Schistosoma alone. CD14 is expressed by the majority of immune response cells - neutrophils, monocytes, macrophages - during the pro-inflammatory response, thus playing a key role in innate immunity and offering, in this case, protection against the severity of the disease relative to the egg load. Indeed, this study also revealed that Schistosoma egg counts were higher in individuals mono-infected with Schistosoma than in co-infected individuals carrying the mutant allele (TT) ^113^. Although the results do not appear to be significant, other studies have shown a link between the number of eggs produced by schistosomes and the intensity of malaria infection during co-infection ^30,31,47,133^. Tokplonou et al. 2023 suggest that eggs may act to activate, suppress or regulate immune pathways.

Other, less obvious factors, such as pathogen species, microgeographical variations ^29^ or the age of individuals may also come into play to explain the high rates of co-infection found in the meta-analysis. Age, in particular, is a recurring factor, with children being more affected by polyparasitism in general ^75,134^ and by schistosomiasis-malaria co-infection ^40,51,102,135^. The meta-analysis carried out between the sexes showed that there were no differences in co-infection rates between men and women, even though many studies report a different prevalence of infection between the two sexes for the two parasites ^64–66,69,71^.

## Limitations

This systematic review did not escape the limitations identified in the studies considered in the meta-analysis. Most of these studies were cross-sectional, which made it difficult to establish conclusively the prevalence of malaria-schistosomiasis co-infections over time. Some studies had deliberately selected infected or co-infected candidates, causing selection bias, as did the refusal of a proportion of the population to participate in certain studies. The potential for co-infection could also vary from one study to another depending on seasonality. Biases may have been introduced because the studies were conducted in very diverse populations, with significant variations in design and implementation methodologies. The sample size was sometimes too small to obtain a significant effect on the prevalence of co-infection and the effects on the immune system, for example. Some papers focused only on asymptomatic or symptomatic forms of malaria, preventing the detection of possible associations with the severity of infection and masking the burden of co-infection. There is no absolute method for detecting Schistosoma or Plasmodium infections, which explains the variations in diagnostic methods used from one study to another. Some of these methods, although considered standard (such as PCR for Plasmodium, Kato-Katz or urine filtration for Schistosoma), may underestimate the prevalence of infection due to their lack of accuracy. However, this systematic review, accompanied by a meta-analysis, has enabled us to update our knowledge on Schistosoma-Plasmodium co-infection and explore the potential interactions between these two parasites, thanks to a comprehensive synthesis of the data collected.

## Conclusion

In conclusion, we have shown that the prevalence of malaria-schistosomiasis co-infection is higher than expected in endemic countries in sub-Saharan Africa. The nature of the interactions between helminths, and more specifically Schistosoma and Plasmodium, during co-infection is still poorly understood, but several relevant avenues are being explored. Knowledge of genetic variation and diversity, parasite biology, population dynamics and parasite transmission and molecular evolution is particularly important for understanding the physiological and immune mechanisms that govern interactions between these two parasites and their hosts. In addition, studies of disease phenotypes in a mouse model or other animal models of co-infection under controlled laboratory conditions, combined with parasite genetics and genomics, can shed light on the pathogenesis of these two parasites and provide new insights into the control and management of disease in humans.

## Data availability

The data adopted in this meta-analysis are available from the corresponding author on reasonable request.

## Acknowledgements

We would like to thank all the authors of the studies included in this systematic review and meta-analysis for their valuable contributions in this field. We are also grateful to our colleagues at the IHPE laboratory in Perpignan for their insightful discussions and technical support during the preparation of this manuscript.

## Contributions

Célia Koellsch, Jérôme Boissier, and Ronaldo de Carvalho Augusto designed the study. Célia Koellsch conducted the database searches, data extraction, and study quality assessment. Célia Koellsch, Jérôme Boissier, and Ronaldo de Carvalho Augusto performed the statistical analyses and interpreted the results. Célia Koellsch wrote the manuscript. Jérôme Boissier and Ronaldo de Carvalho Augusto reviewed the manuscript. All authors approved the submission of the manuscript.

## Corresponding authors

Correspondence to Ronaldo de Carvalho Augusto

## Appendixes

### Appendix S1

Checklist for PRISMA 2020

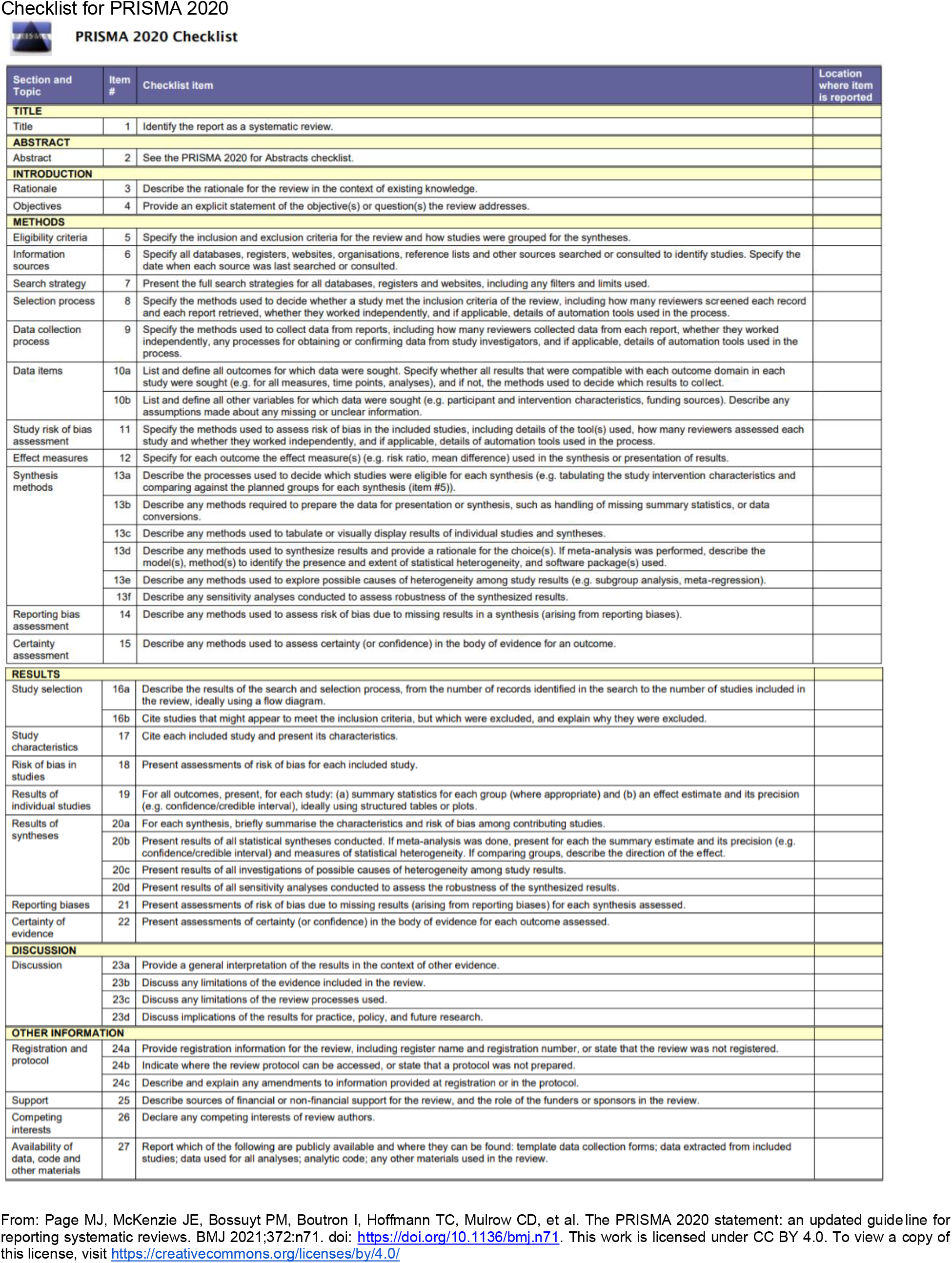

### Appendix S2

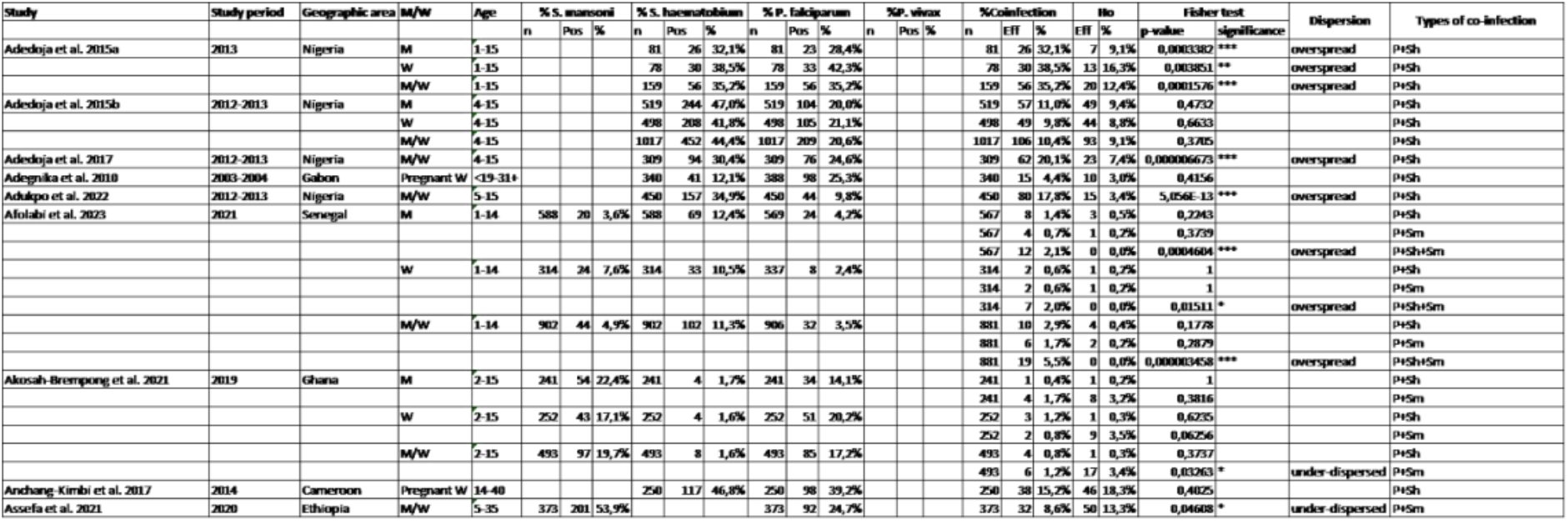

### Appendix S3

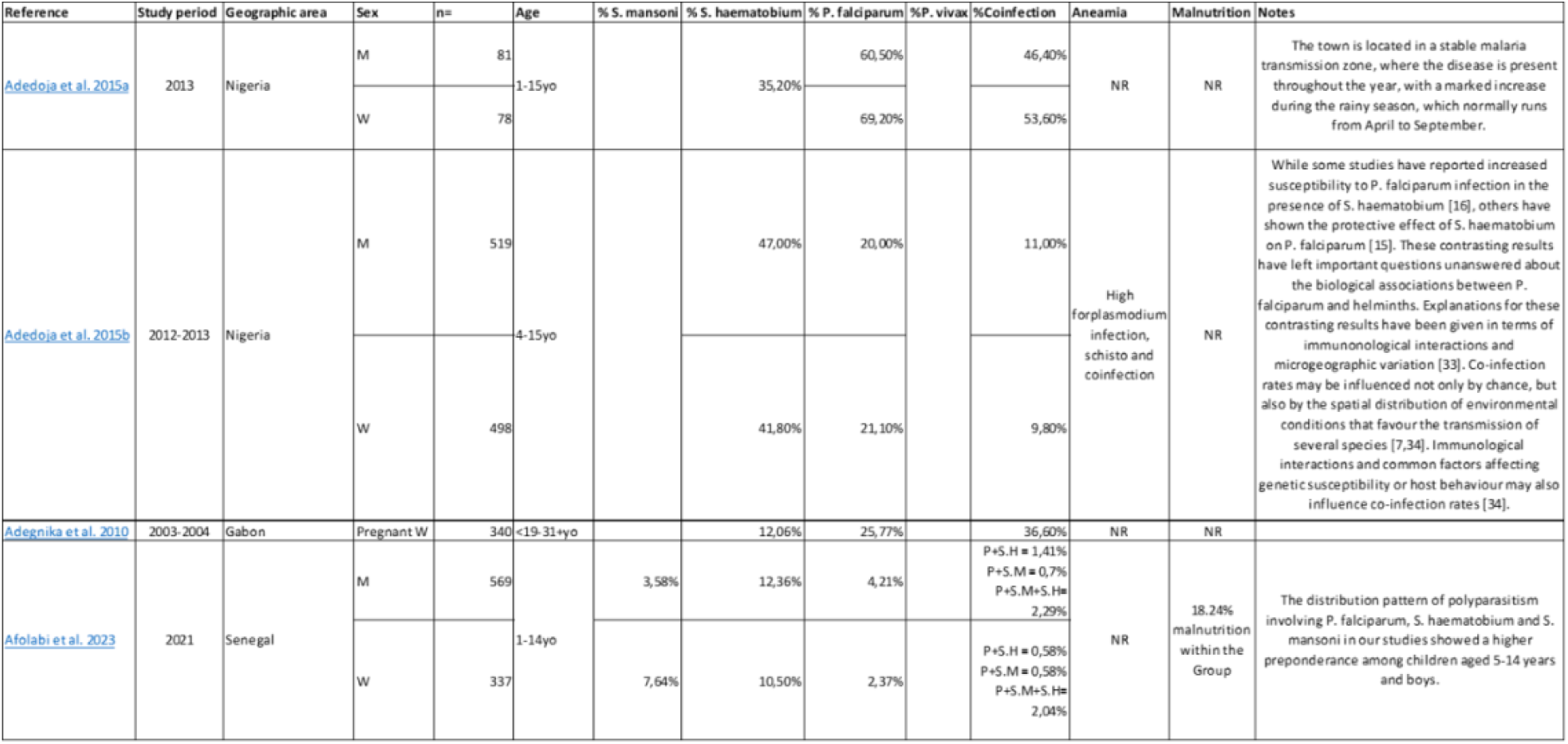

## Supplementary data

**Supplementary Figure 1.**
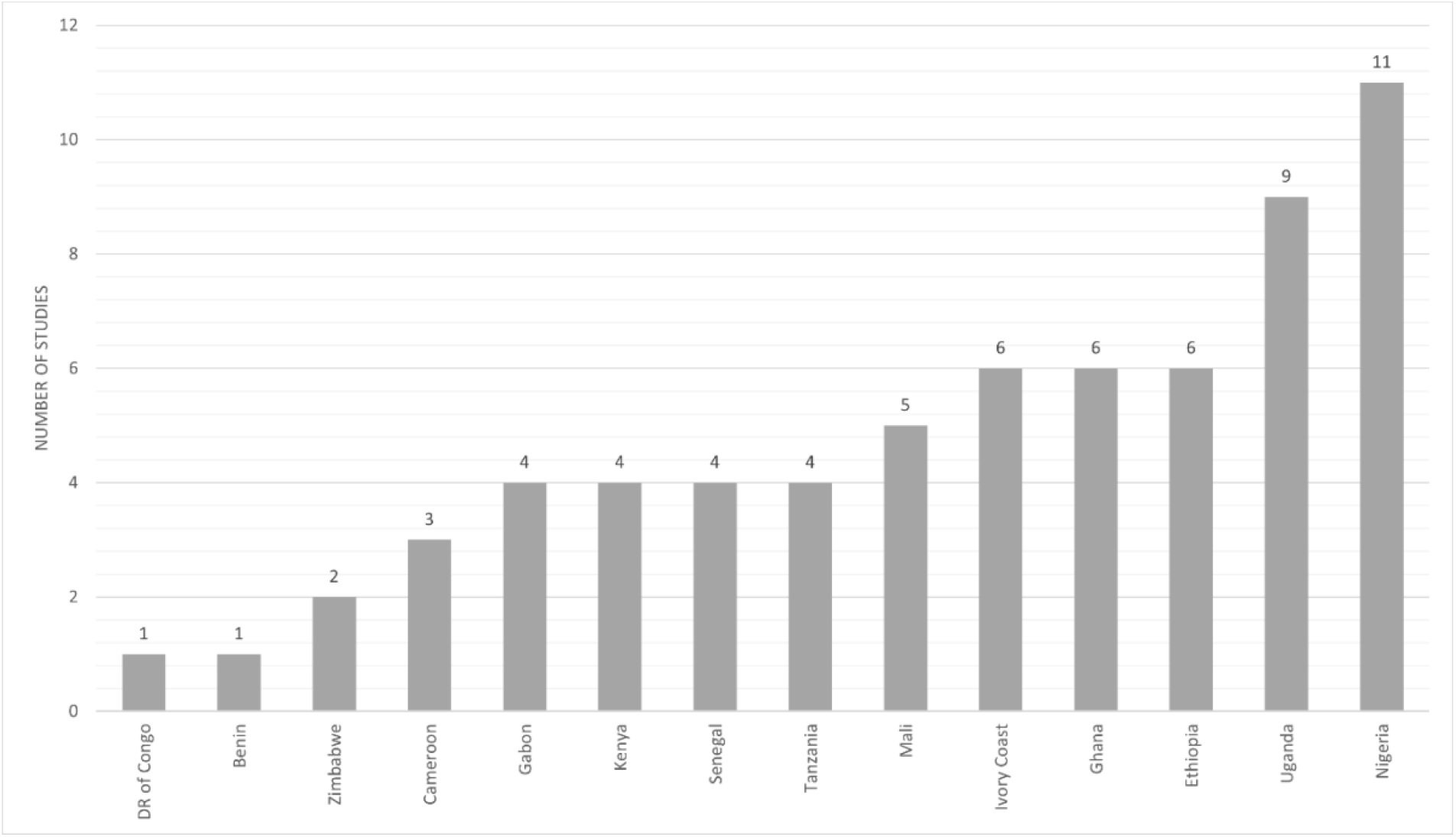
Geographical areas most frequently sampled for the prevalence of co-infection. This bar chart illustrates the number of studies included in the systematic review and meta-analysis according to the sampled countries. Nigeria reported the highest number of studies (n = 11), followed by Uganda (n = 9), Ethiopia (n = 6), Ghana (n = 6), and Ivory Coast (n = 6). Mali (n = 5), Tanzania (n = 4), Senegal (n = 4), Kenya (n = 4), Gabon (n = 4), and Cameroon (n = 3) follow. Zimbabwe (n = 2), Benin (n = 1), and the Democratic Republic of Congo (n = 1) contributed the fewest studies. Overall, the figure highlights the geographical heterogeneity of the studies, with a predominance in West and East African countries where co-infections of Plasmodium spp. and Schistosoma spp. are most frequently investigated

**Supplementary Figure 2.**
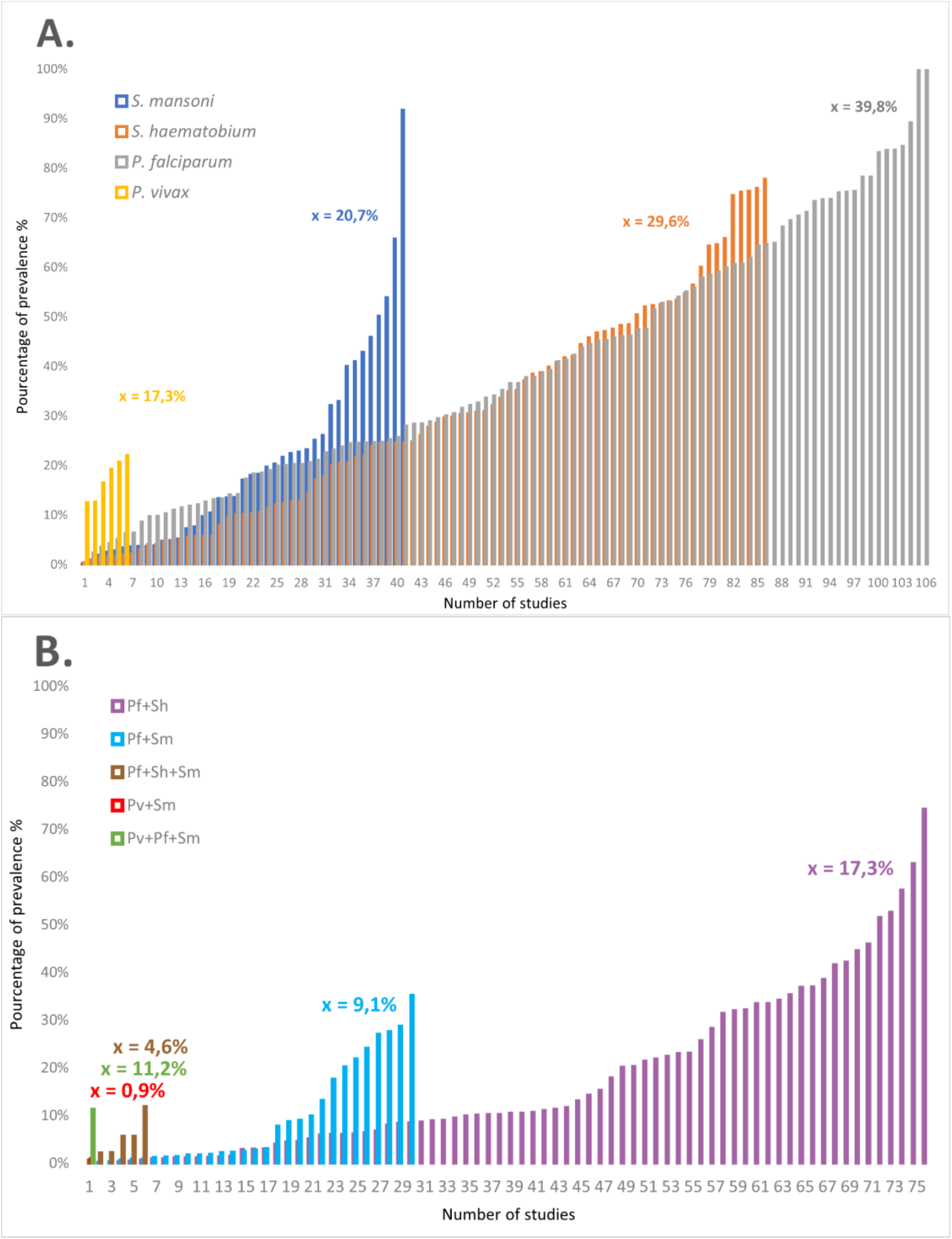
A. Prevalences of single infections and B. co-infections of the two species of Schistosoma (S. mansoni and S. haematobium) and the two species of Plasmodium (P. falciparum and P. vivax) in integrated studies for epidemiological data and meta-analysis. The averages for each prevalence - single and co-infections - are shown in the colour associated with the legend. The y-axis shows the percentage prevalence of single and co-infections, while the x-axis shows the number of prevalences found in the total of 59 studies analysed (several prevalences can be found in the same article).

**Supplementary Figure 3.**
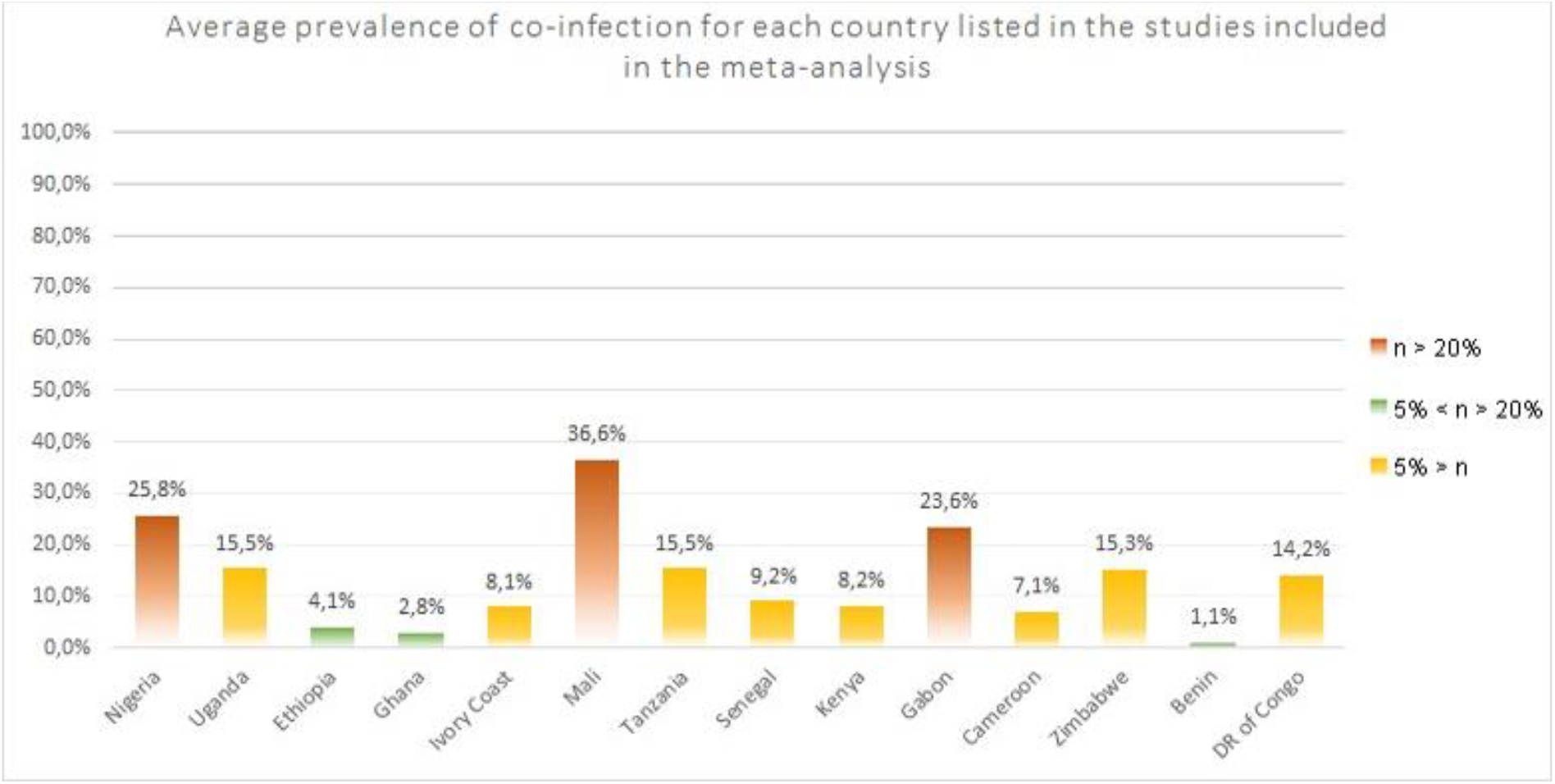
Average prevalence of co-infection for each country listed in the studies included in the meta-analysis. This figure illustrates the average prevalence of Plasmodium–Schistosoma co-infections reported in the studies included in the meta-analysis, showing marked geographical variation across African countries. The highest prevalence rates were observed in Mali (36.6%), Nigeria (25.8%), and Gabon (23.6%), suggesting possible hotspots of co-infection. Intermediate prevalence levels were found in Uganda (15.5%), Tanzania (15.5%), Senegal (9.2%), Kenya (8.2%), and Ivory Coast (8.1%). In contrast, lower co-infection rates were reported in Ethiopia (4.1%), Ghana (2.8%), and Benin (1.1%). These findings highlight substantial geographical heterogeneity in the burden of Plasmodium–Schistosoma co-infections across sub-Saharan Africa.

***Supplementary Table 1. Prevalence and distribution of co-infections between schistosomiasis and malaria reported in various African studies***. *This table summarises data from several epidemiological studies conducted in Africa and included in the meta-analysis on the prevalence of infections with Schistosoma mansoni, Schistosoma haematobium, Plasmodium falciparum and Plasmodium vivax, as well as their co-infections. The information includes study periods, geographical areas, demographic characteristics of the populations studied, infection rates, types of co-infection observed, and associated statistical results (Fisher’s exact test, dispersion)*.

***Supplementary Table 2. Epidemiological surveys on the co-occurrence of schistosomiasis and malaria infections in Africa***. *This table presents a summary of cross-sectional studies reporting the prevalence of Schistosoma mansoni, Schistosoma haematobium, Plasmodium falciparum and Plasmodium vivax infections, as well as their co-infections. The data include the characteristics of the populations studied (sample size, age, sex, geographical area), the presence of anaemia or malnutrition, and the main methodological limitations reported by the authors. Taken together, the data highlight the epidemiological variability and contextual factors influencing the association between these parasitic diseases*.

***Supplementary Table 3. Functional studies on the immunological and biochemical impact of schistosomiasis– malaria co-infections***. *This table summarises functional studies conducted in Africa exploring the immunological, biochemical and clinical consequences of co-infections between Schistosoma spp. and Plasmodium spp. The data include the characteristics of the populations studied, the prevalence of single and combined infections, and the observed effects on immune responses (cytokines, antibodies, regulatory cells), biochemical parameters (ALT, AST, bilirubin, glucose, proteins), and clinical markers (anaemia, malnutrition). The reported studies highlight complex interactions between the two parasitic diseases, which may modulate susceptibility, vaccine response, and the clinical severity of malaria*.

